# Parkinson’s Disease, Smoking, and Lower Endoscopy

**DOI:** 10.1101/2020.11.10.20229450

**Authors:** Susan M. Davis

## Abstract

Parkinson’s disease (PD) is inversely associated with smoking. Whether this association is due to a causal relationship or to confounding by a covariate of smoking is still debated.

The Institute for Health Metrics and Evaluation (IHME) released refreshed data on October 15, 2020. This study included that recently released data. The study included populations of the United States and ten U.S. states from 2004 to 2018. The ten U.S. states included the five states with the highest PD incidence rates in 2019 (Maine, Vermont, Kansas, Alaska, Missouri) and the five states with the lowest PD incidence rates in 2019 (Arkansas, Mississippi, South Dakota, Nebraska, Delaware). The study used scatter plots to explore the association between PD incidence and smoking and the association between PD incidence and a covariate of smoking, lower endoscopy utilization.

For PD verses smoking, the results indicate that there is an inverse correlation for the United States, but there is no association for the ten states. The coefficient of determination (R^2^) for the United States was 0.714 and ranged from a low of 0.004 for South Dakota to 0.613 for Mississippi. The average R ^2^ for the ten states was 0.357.

For PD verses lower endoscopy, the results indicate that the best model fit to the data is a polynomial. When the fitting curve examined in the regression analysis was a 3rd order (cubic) polynomial, there was a positive correlation between PD and lower endoscopy for the United States and for all ten states. The R^2^ for the US was 0.971 and ranged from a low of 0.709 for Alaska to 0.970 for Kansas. The average R^2^ for the ten states was 0.878.

The results suggest that the inverse association between PD incidence and smoking is confounded by a positive association between PD and lower endoscopy utilization. Further investigation of a possible relationship between PD incidence and lower endoscopy utilization is warranted and may provide a means for reducing PD incidence.

## INTRODUCTION

Parkinson’s disease (PD) incidence rates have been increasing [1]. There is no known cause for most cases of PD, and they are thought to result from a complex combination of many factors, including industrialization, environmental toxins, lifestyle, genetics, and age [2-3].

PD is related to smoking, with current smokers having a lower risk for PD when compared to never smokers [2-4]. Of eighty-four risk factors explored by the Global Burden of Diseases, Injuries, and Risk Factors (GBD), smoking was the only risk factor deemed to have a relationship with PD [2]. The inverse relationship between PD and smoking may be causal or it may be due to confounding by a covariate of smoking.

One covariate of smoking is colorectal cancer (CRC) screening. Several studies have found that current smokers are less likely to participate in CRC screening compared to never smokers [5-9]. In the United States, the most commonly used CRC screening test is colonoscopy [10]. Compared with never smokers, current smokers are less likely to ever have received a colonoscopy [5].

Colonoscopy is a lower endoscopic procedure that examines the entire colon, and precancerous polyps can be removed during the procedure, thereby reducing CRC risk [11-12]. Another lower endoscopic procedure used for CRC screening is sigmoidoscopy, which is similar to colonoscopy, but examines only a portion of the colon [12-13].

A previous study that explored the association between PD incidence and smoking and the association between PD incidence and lower endoscopy utilization posted on MedRxiv on September 23, 2020 (https://medrxiv.org/cgi/content/short/2020.09.20.20198390v1). That study included data obtained from the Institute for Health Metrics and Evaluation (IHME)’s Global Burden of Diseases, Injuries, and Risk Factors (GBD) Compare visualization system on September 15, 2020. (Data used in that study is now available at https://gbd2017.healthdata.org/gbd-compare/). That study concluded that the inverse association between PD incidence and smoking is confounded by a direct association between PD and lower endoscopy utilization.

On October 15, 2020, the IHME refreshed the data to reflect the new results from the GBD 2019 (available at https://vizhub.healthdata.org/gbd-compare). The present study included this refreshed data.

## METHODS

The present study comprises scatter plots of PD incidence versus current smoking and PD incidence versus lower endoscopy utilization for different populations.

This study included populations of the United States (all 50 states and the District of Columbia) and ten U.S. states from 2004 to 2018. The ten U.S. states included the five states with the highest PD incidence rates in 2019 (Maine, Vermont, Kansas, Alaska, Missouri) and the five states with the lowest PD incidence rates in 2019 (Arkansas, Mississippi, South Dakota, Nebraska, Delaware). Compared to the previous study, Colorado was replaced by Alaska in the group of five states with the highest PD incidence rates, and Alabama was replaced by Delaware in the group of five states with the lowest PD incidence rates.

PD incidence rates (age standardized, new cases per 100,000) were obtained from the Institute for Health Metrics and Evaluation (IHME)’s Global Burden of Diseases, Injuries, and Risk Factors (GBD) Compare visualization system (available at https://vizhub.healthdata.org/gbd-compare) [14]. Methodology, sample size, and 95% confidence intervals may be obtained from the IHME.

Rates of current smoking (% adults who are current smokers) were obtained from the Centers for Disease Control and Prevention (CDC)’s Behavioral Risk Factor Surveillance System (BRFSS) Web Enabled Analysis Tool (WEAT) (available at https://www.cdc.gov/brfss/data_tools.htm) [15]. For 2004, the column variable for current smoking was “Tobacco Use: Calculated variable for adults who are current smokers (_RFSMOK2)”. For all other years, the column variable for current smoking was “Tobacco Use: Calculated variable for adults who are current smokers (_RFSMOK3)”. Methodology, sample size, and 95% confidence intervals may be obtained from the CDC.

Rates of lower endoscopy utilization (% who ever had a sigmoidoscopy/colonoscopy) were obtained from the Centers for Disease Control and Prevention (CDC)’s Behavioral Risk Factor Surveillance System (BRFSS) Web Enabled Analysis Tool (WEAT) (available at https://www.cdc.gov/brfss/data_tools.htm) [15]. The column variable for lower endoscopy utilization was “Colorectal Cancer Screening: Ever had a sigmoidoscopy/colonoscopy (HADSIGM3)”. Methodology, sample size, and 95% confidence intervals may be obtained from the CDC.

This study included the years 2004, 2006, 2008, 2010, 2012, 2014, 2016 and 2018, as those were the only years for which lower endoscopy utilization data were available from WEAT. The scatter plots, trendline equations and coefficients of determination (R^2^) were generated using Microsoft Excel.

## RESULTS

Figures 1a and 1b are scatter plots of PD verses smoking. Figure 1a shows the five states with the highest PD incidence rates in 2019 and the US. Figure 1b shows the five states with the lowest PD incidence rates in 2019 and the US.

**Figure 1a:**
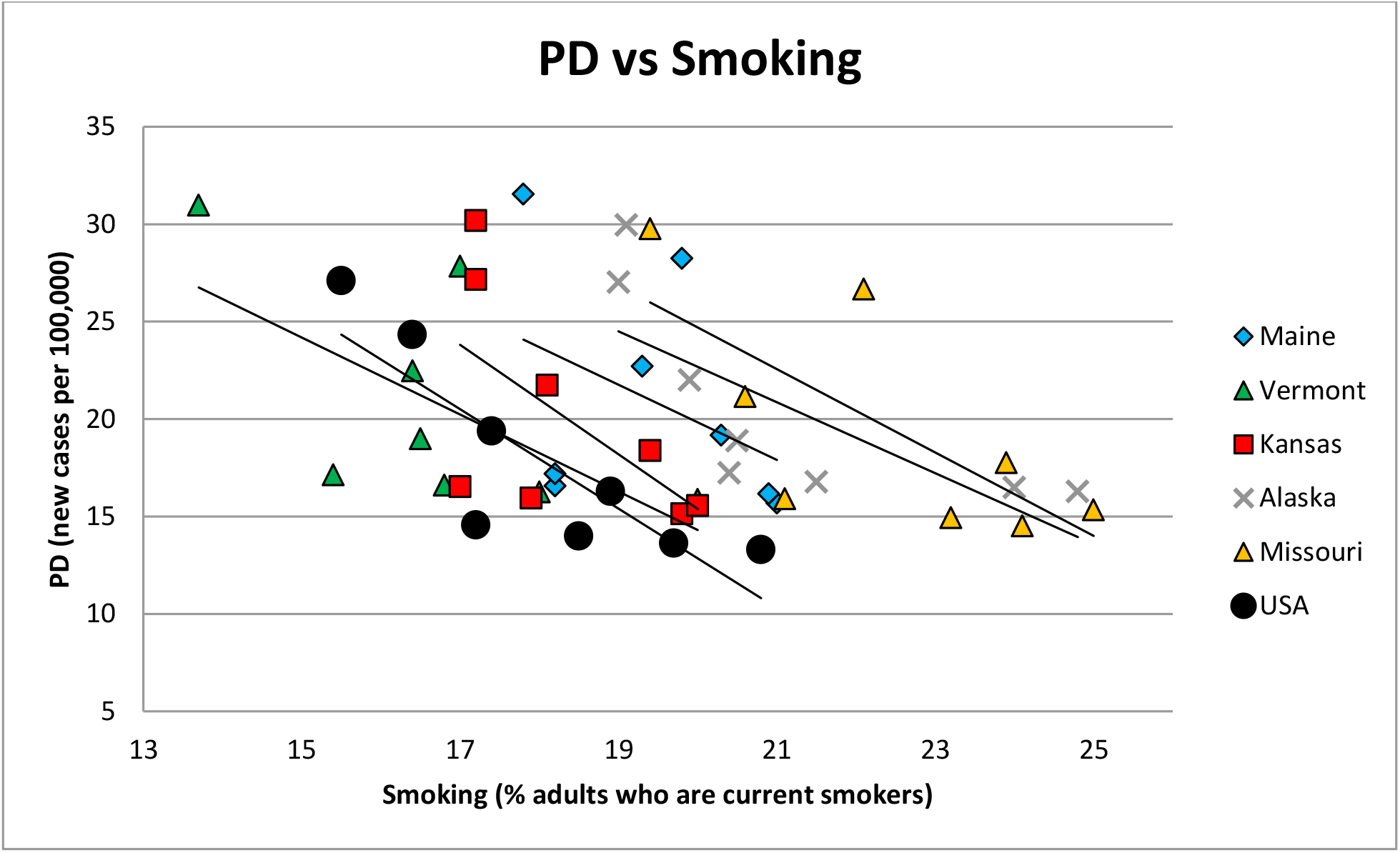
PD vs smoking for the five states with the highest PD incidence rates and the US.

**Figure 1b:**
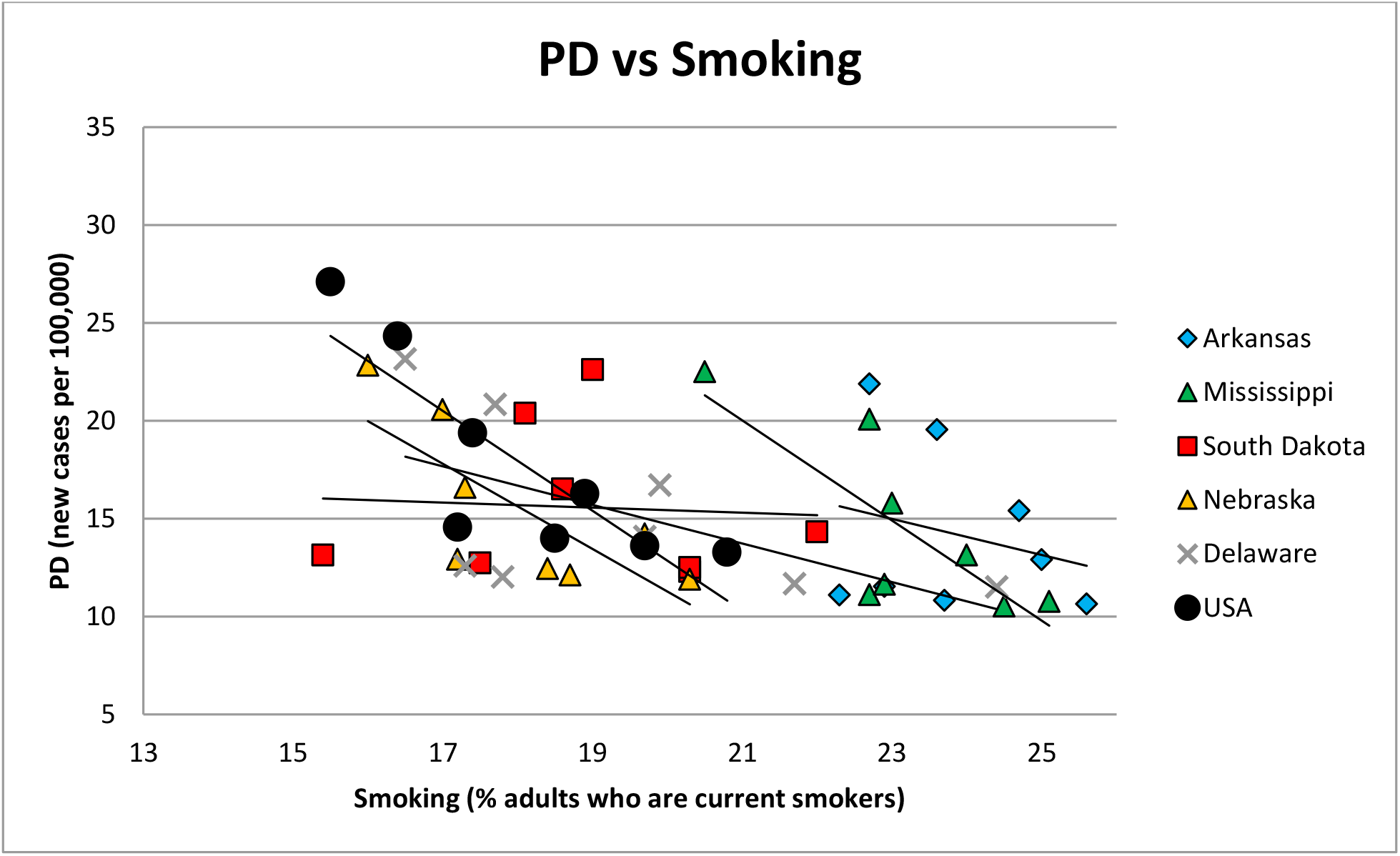
PD vs smoking for the five states with the lowest PD incidence rates and the US.

Figures 2a and 2b are scatter plots of PD verses lower endoscopy. Figure 2a shows the five states with the highest PD incidence rates in 2019 and the US. Figure 2b shows the five states with the lowest PD incidence rates in 2019 and the US.

**Figure 2a:**
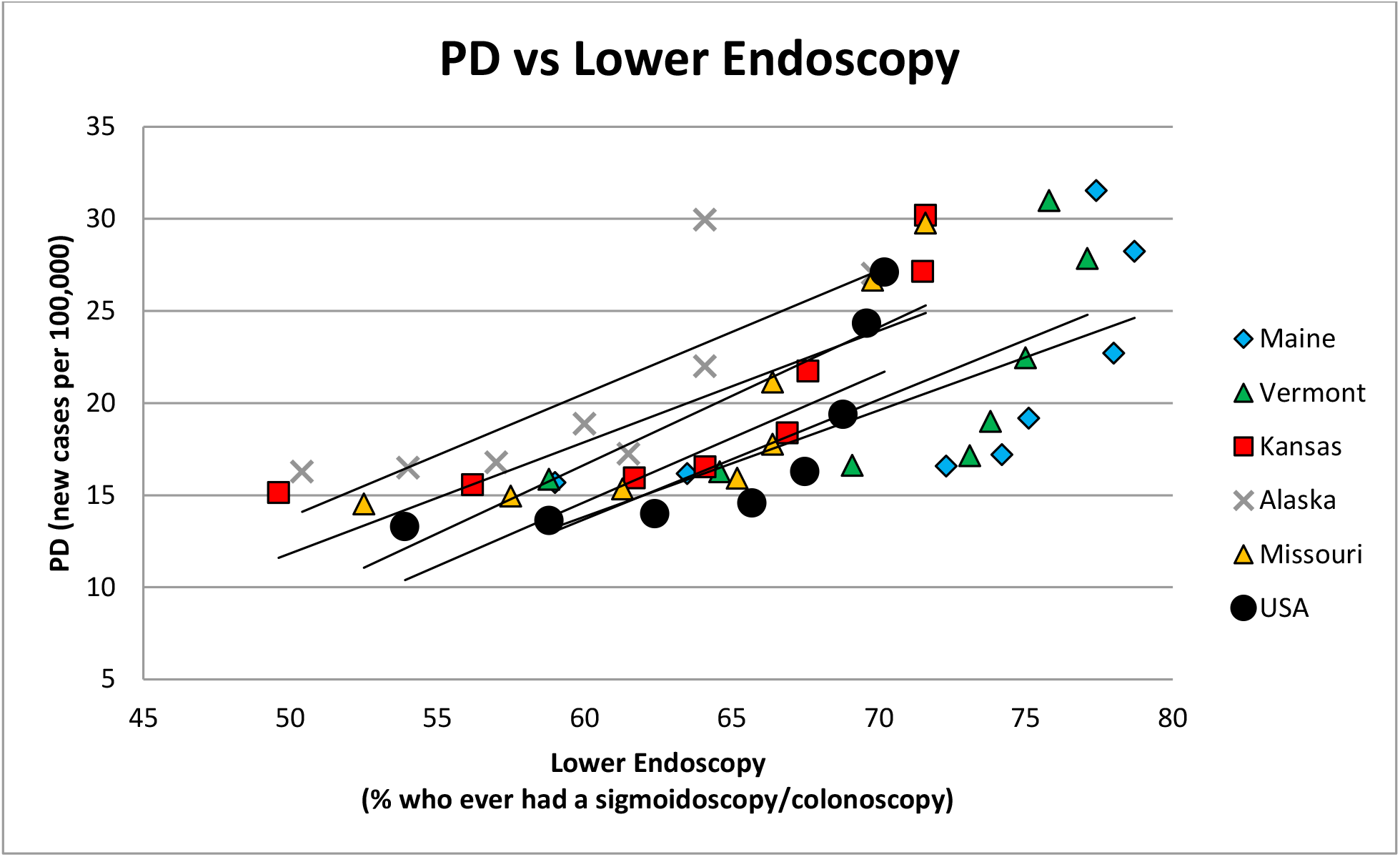
PD vs lower endoscopy for the five states with the highest PD incidence rates and the US.

**Figure 2b:**
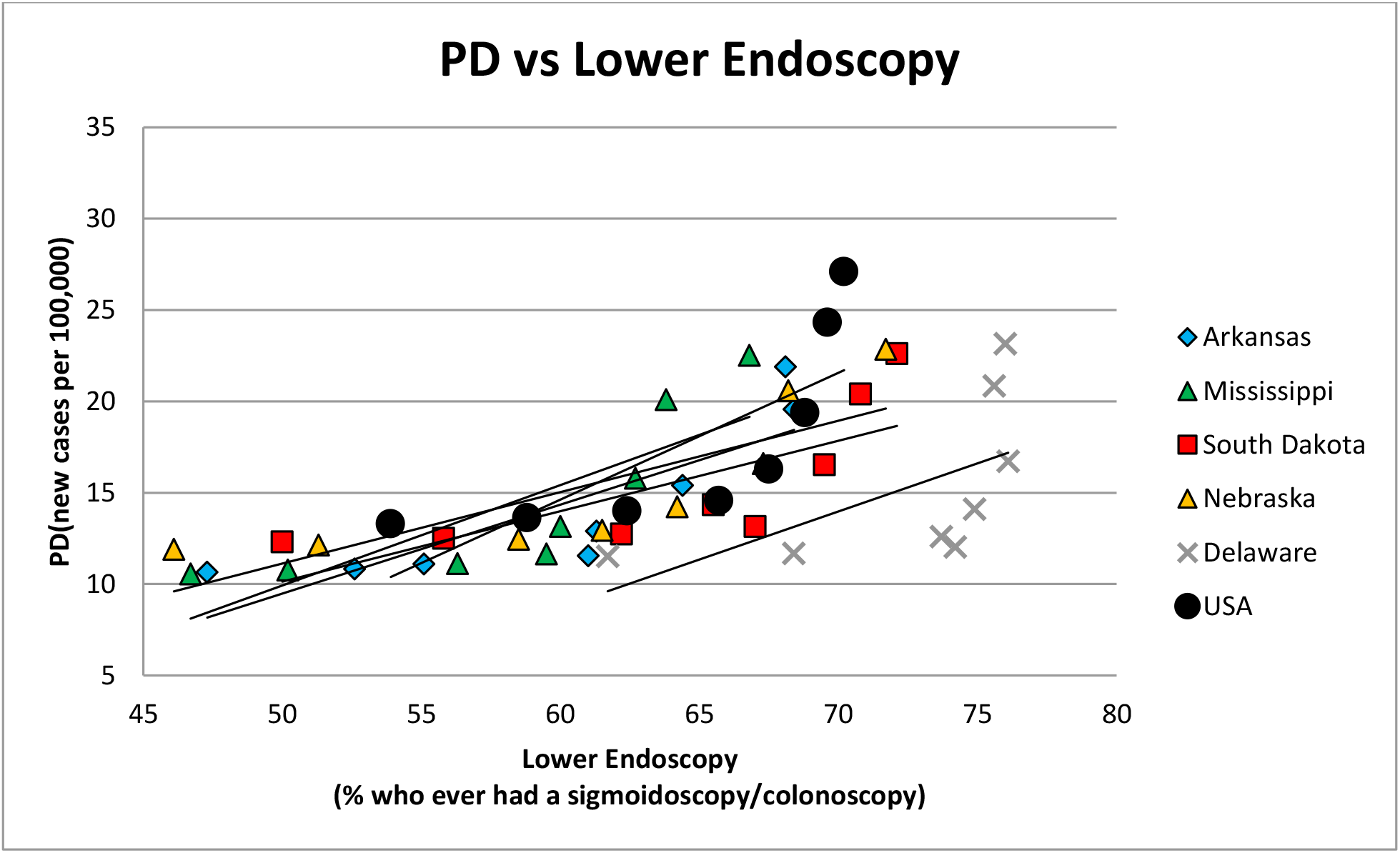
PD vs lower endoscopy for the five states with the lowest PD incidence rates and the US.

Table 1 shows the data plotted in the figures, and the trendline equations and R^2^ values for linear fitting curves. Tables 2-4 show the trendline equations and R^2^ values for 2nd, 3rd and 4th order polynomial fitting curves, respectively.

**Table 1:**
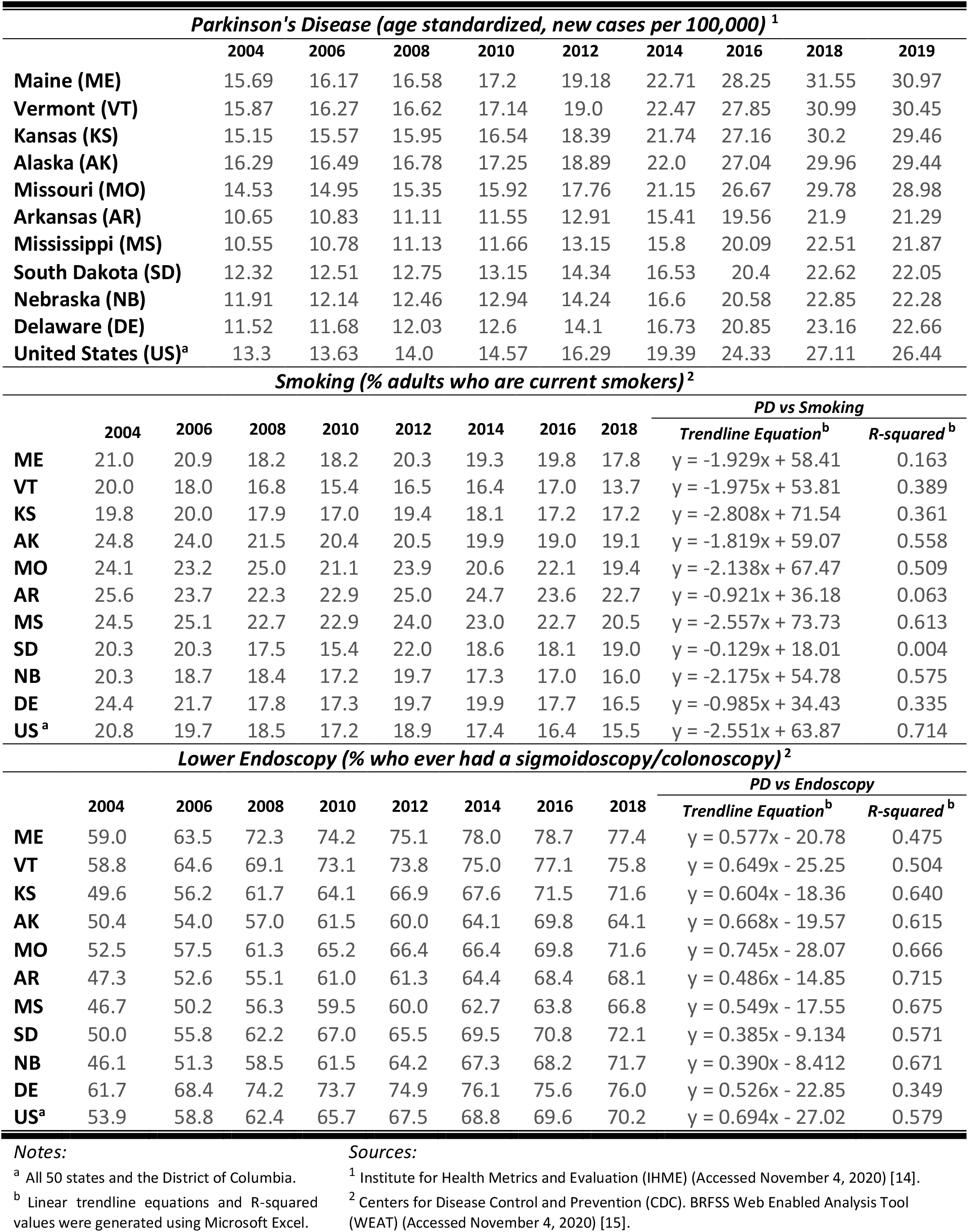
Data plotted in Figures 1a,b and 2a,b; trendline equations and R^2^ values for linear fitting curves.

| Parkinson's Disease (age standardized, new cases per 100,000) <sup>1</sup> |  |  |  |  |  |  |  |  |  |  |
| --- | --- | --- | --- | --- | --- | --- | --- | --- | --- | --- |
|  | 2004 | 2006 | 2008 | 2010 | 2012 | 2014 | 2016 | 2018 | 2019 |  |
| Maine (ME) | 15.69 | 16.17 | 16.58 | 17.2 | 19.18 | 22.71 | 28.25 | 31.55 | 30.97 |  |
| Vermont (VT) | 15.87 | 16.27 | 16.62 | 17.14 | 19.0 | 22.47 | 27.85 | 30.99 | 30.45 |  |
| Kansas (KS) | 15.15 | 15.57 | 15.95 | 16.54 | 18.39 | 21.74 | 27.16 | 30.2 | 29.46 |  |
| Alaska (AK) | 16.29 | 16.49 | 16.78 | 17.25 | 18.89 | 22.0 | 27.04 | 29.96 | 29.44 |  |
| Missouri (MO) | 14.53 | 14.95 | 15.35 | 15.92 | 17.76 | 21.15 | 26.67 | 29.78 | 28.98 |  |
| Arkansas (AR) | 10.65 | 10.83 | 11.11 | 11.55 | 12.91 | 15.41 | 19.56 | 21.9 | 21.29 |  |
| Mississippi (MS) | 10.55 | 10.78 | 11.13 | 11.66 | 13.15 | 15.8 | 20.09 | 22.51 | 21.87 |  |
| South Dakota (SD) | 12.32 | 12.51 | 12.75 | 13.15 | 14.34 | 16.53 | 20.4 | 22.62 | 22.05 |  |
| Nebraska (NB) | 11.91 | 12.14 | 12.46 | 12.94 | 14.24 | 16.6 | 20.58 | 22.85 | 22.28 |  |
| Delaware (DE) | 11.52 | 11.68 | 12.03 | 12.6 | 14.1 | 16.73 | 20.85 | 23.16 | 22.66 |  |
| United States (US)<sup>a</sup> | 13.3 | 13.63 | 14.0 | 14.57 | 16.29 | 19.39 | 24.33 | 27.11 | 26.44 |  |
| Smoking (% adults who are current smokers)<sup>2</sup> |  |  |  |  |  |  |  |  |  |  |
|  | 2004 | 2006 | 2008 | 2010 | 2012 | 2014 | 2016 | 2018 | PD vs Smoking |  |
|  |  |  |  |  |  |  |  |  | Trendline Equation<sup>b</sup> | R-squared<sup>b</sup> |
| ME | 21.0 | 20.9 | 18.2 | 18.2 | 20.3 | 19.3 | 19.8 | 17.8 | y = -1.929x + 58.41 | 0.163 |
| VT | 20.0 | 18.0 | 16.8 | 15.4 | 16.5 | 16.4 | 17.0 | 13.7 | y = -1.975x + 53.81 | 0.389 |
| KS | 19.8 | 20.0 | 17.9 | 17.0 | 19.4 | 18.1 | 17.2 | 17.2 | y = -2.808x + 71.54 | 0.361 |
| AK | 24.8 | 24.0 | 21.5 | 20.4 | 20.5 | 19.9 | 19.0 | 19.1 | y = -1.819x + 59.07 | 0.558 |
| MO | 24.1 | 23.2 | 25.0 | 21.1 | 23.9 | 20.6 | 22.1 | 19.4 | y = -2.138x + 67.47 | 0.509 |
| AR | 25.6 | 23.7 | 22.3 | 22.9 | 25.0 | 24.7 | 23.6 | 22.7 | y = -0.921x + 36.18 | 0.063 |
| MS | 24.5 | 25.1 | 22.7 | 22.9 | 24.0 | 23.0 | 22.7 | 20.5 | y = -2.557x + 73.73 | 0.613 |
| SD | 20.3 | 20.3 | 17.5 | 15.4 | 22.0 | 18.6 | 18.1 | 19.0 | y = -0.129x + 18.01 | 0.004 |
| NB | 20.3 | 18.7 | 18.4 | 17.2 | 19.7 | 17.3 | 17.0 | 16.0 | y = -2.175x + 54.78 | 0.575 |
| DE | 24.4 | 21.7 | 17.8 | 17.3 | 19.7 | 19.9 | 17.7 | 16.5 | y = -0.985x + 34.43 | 0.335 |
| US<sup>a</sup> | 20.8 | 19.7 | 18.5 | 17.2 | 18.9 | 17.4 | 16.4 | 15.5 | y = -2.551x + 63.87 | 0.714 |
| Lower Endoscopy (% who ever had a sigmoidoscopy/colonoscopy)<sup>2</sup> |  |  |  |  |  |  |  |  |  |  |
|  | 2004 | 2006 | 2008 | 2010 | 2012 | 2014 | 2016 | 2018 | PD vs Endoscopy |  |
|  |  |  |  |  |  |  |  |  | Trendline Equation<sup>b</sup> | R-squared<sup>b</sup> |
| ME | 59.0 | 63.5 | 72.3 | 74.2 | 75.1 | 78.0 | 78.7 | 77.4 | y = 0.577x - 20.78 | 0.475 |
| VT | 58.8 | 64.6 | 69.1 | 73.1 | 73.8 | 75.0 | 77.1 | 75.8 | y = 0.649x - 25.25 | 0.504 |
| KS | 49.6 | 56.2 | 61.7 | 64.1 | 66.9 | 67.6 | 71.5 | 71.6 | y = 0.604x - 18.36 | 0.640 |
| AK | 50.4 | 54.0 | 57.0 | 61.5 | 60.0 | 64.1 | 69.8 | 64.1 | y = 0.668x - 19.57 | 0.615 |
| MO | 52.5 | 57.5 | 61.3 | 65.2 | 66.4 | 66.4 | 69.8 | 71.6 | y = 0.745x - 28.07 | 0.666 |
| AR | 47.3 | 52.6 | 55.1 | 61.0 | 61.3 | 64.4 | 68.4 | 68.1 | y = 0.486x - 14.85 | 0.715 |
| MS | 46.7 | 50.2 | 56.3 | 59.5 | 60.0 | 62.7 | 63.8 | 66.8 | y = 0.549x - 17.55 | 0.675 |
| SD | 50.0 | 55.8 | 62.2 | 67.0 | 65.5 | 69.5 | 70.8 | 72.1 | y = 0.385x - 9.134 | 0.571 |
| NB | 46.1 | 51.3 | 58.5 | 61.5 | 64.2 | 67.3 | 68.2 | 71.7 | y = 0.390x - 8.412 | 0.671 |
| DE | 61.7 | 68.4 | 74.2 | 73.7 | 74.9 | 76.1 | 75.6 | 76.0 | y = 0.526x - 22.85 | 0.349 |
| US<sup>a</sup> | 53.9 | 58.8 | 62.4 | 65.7 | 67.5 | 68.8 | 69.6 | 70.2 | y = 0.694x - 27.02 | 0.579 |

**Notes:**
<sup>a</sup> All 50 states and the District of Columbia.
<sup>b</sup> Linear trendline equations and R-squared values were generated using Microsoft Excel.
**Sources:**
<sup>1</sup> Institute for Health Metrics and Evaluation (IHME) (Accessed November 4, 2020) [14].
<sup>2</sup> Centers for Disease Control and Prevention (CDC). BRFSS Web Enabled Analysis Tool (WEAT) (Accessed November 4, 2020) [15].

**Table 2:** Trendline equations and R^2^ values for 2nd order polynomial fitting curves.

| **2nd Order Polynomial** |  |  |
| --- | --- | --- |
| **PD vs Smoking** |  |  |
|  | **Trendline Equation<sup>b</sup>** | **R-squared<sup>b</sup>** |
| **Maine** | $y = -1.119x^2 + 41.51x - 361.7$ | 0.198 |
| **Vermont** | $y = 0.314x^2 - 12.58x + 142.3$ | 0.445 |
| **Kansas** | $y = 0.086x^2 - 5.990x + 100.8$ | 0.361 |
| **Alaska** | $y = 0.970x^2 - 44.37x + 520.8$ | 0.906 |
| **Missouri** | $y = 0.366x^2 - 18.42x + 247.1$ | 0.545 |
| **Arkansas** | $y = -1.293x^2 + 61.00x - 703.4$ | 0.160 |
| **Mississippi** | $y = 0.330x^2 - 17.63x + 245.3$ | 0.641 |
| **South Dakota** | $y = -0.387x^2 + 14.38x - 116.3$ | 0.209 |
| **Nebraska** | $y = 0.994x^2 - 38.40x + 382.9$ | 0.764 |
| **Delaware** | $y = 0.152x^2 - 7.195x + 96.62$ | 0.380 |
| **United States<sup>a</sup>** | $y = 0.690x^2 - 27.62x + 289.4$ | 0.851 |
| **PD vs Lower Endoscopy** |  |  |
|  | **Trendline Equation<sup>b</sup>** | **R-squared<sup>b</sup>** |
| **Maine** | $y = 0.087x^2 - 11.50x + 391.0$ | 0.719 |
| **Vermont** | $y = 0.092x^2 - 11.87x + 397.3$ | 0.747 |
| **Kansas** | $y = 0.060x^2 - 6.779x + 203.5$ | 0.931 |
| **Alaska** | $y = 0.025x^2 - 2.339x + 69.60$ | 0.649 |
| **Missouri** | $y = 0.082x^2 - 9.462x + 285.5$ | 0.932 |
| **Arkansas** | $y = 0.043x^2 - 4.621x + 132.0$ | 0.939 |
| **Mississippi** | $y = 0.060x^2 - 6.263x + 172.1$ | 0.942 |
| **South Dakota** | $y = 0.046x^2 - 5.267x + 161.2$ | 0.877 |
| **Nebraska** | $y = 0.033x^2 - 3.568x + 105.4$ | 0.935 |
| **Delaware** | $y = 0.120x^2 - 16.16x + 549.9$ | 0.581 |
| **United States<sup>a</sup>** | $y = 0.106x^2 - 12.55x + 382.7$ | 0.851 |

Notes:
<sup>a</sup> All 50 states and the District of Columbia.
<sup>b</sup> Polynomial trendline equations and R-squared values were generated using Microsoft Excel.

**Table 3:** Trendline equations and R^2^ values for 3rd order polynomial fitting curves.

| **3rd Order Polynomial** |  |  |
| --- | --- | --- |
| **PD vs Smoking** |  |  |
|  | **Trendline Equation<sup>b</sup>** | **R-squared<sup>b</sup>** |
| **Maine** | $y = -5.453x^3 + 317.5x^2 - 6152.x + 39702$ | 0.662 |
| **Vermont** | $y = -0.303x^3 + 15.63x^2 - 267.6x + 1543.$ | 0.528 |
| **Kansas** | $y = 0.646x^3 - 35.64x^2 + 651x - 3918.$ | 0.366 |
| **Alaska** | $y = -0.225x^3 + 15.60x^2 - 359.2x + 2768.$ | 0.941 |
| **Missouri** | $y = -0.221x^3 + 15.09x^2 - 343.8x + 2633.$ | 0.582 |
| **Arkansas** | $y = 0.390x^3 - 29.35x^2 + 731.8x - 6042.$ | 0.167 |
| **Mississippi** | $y = -0.024x^3 + 2.023x^2 - 56.12x + 535.9$ | 0.641 |
| **South Dakota** | $y = 0.125x^3 - 7.426x^2 + 144.7x - 914.3$ | 0.266 |
| **Nebraska** | $y = -0.132x^3 + 8.205x^2 - 168.5x + 1162.$ | 0.769 |
| **Delaware** | $y = -0.089x^3 + 5.663x^2 - 118.7x + 841.5.$ | 0.431 |
| **United States <sup>a</sup>** | $y = -0.103x^3 + 6.336x^2 - 129.5x + 899.3$ | 0.858 |
| **PD vs Lower Endoscopy** |  |  |
|  | **Trendline Equation<sup>b</sup>** | **R-squared<sup>b</sup>** |
| **Maine** | $y = 0.006x^3 - 1.180x^2 + 76.12x - 1615.$ | 0.758 |
| **Vermont** | $y = 0.010x^3 - 2.007x^2 + 130.2x - 2791.$ | 0.819 |
| **Kansas** | $y = 0.004x^3 - 0.684x^2 + 38.32x - 697.9$ | 0.970 |
| **Alaska** | $y = -0.006x^3 + 1.203x^2 - 72.94x + 1469.$ | 0.709 |
| **Missouri** | $y = 0.004x^3 - 0.762x^2 + 42.81x - 786.1$ | 0.953 |
| **Arkansas** | $y = 0.002x^3 - 0.372x^2 + 19.38x - 324.8$ | 0.960 |
| **Mississippi** | $y = 0.001x^3 - 0.200x^2 + 8.515x - 104.7$ | 0.947 |
| **South Dakota** | $y = 0.004x^3 - 0.717x^2 + 41.21x - 772.7$ | 0.959 |
| **Nebraska** | $y = 0.001x^3 - 0.177x^2 + 8.774x - 132.0$ | 0.950 |
| **Delaware** | $y = 0.032x^3 - 6.682x^2 + 452.6x - 10182$ | 0.753 |
| **United States <sup>a</sup>** | $y = 0.017x^3 - 3.116x^2 + 186.7x - 3704$ | 0.971 |

Notes:
<sup>a</sup> All 50 states and the District of Columbia.
<sup>b</sup> Polynomial trendline equations and R-squared values were generated using Microsoft Excel.

**Table 4:** Trendline equations and R^2^ values for 4th order polynomial fitting curves.

| **4th Order Polynomial** |  |  |
| --- | --- | --- |
| **PD vs Smoking** |  |  |
|  | **Trendline Equation<sup>b</sup>** | **R-squared<sup>b</sup>** |
| **Maine** | $y = 4.722x^4 - 371.0x^3 + 10917x^2 - 14257x + 69732$ | 0.949 |
| **Vermont** | $y = 0.381x^4 - 25.96x^3 + 658.3x^2 - 7380.x + 30876$ | 0.689 |
| **Kansas** | $y = -7.587x^4 + 564.3x^3 - 15723x^2 + 19442x - 90041$ | 0.639 |
| **Alaska** | $y = 0.005x^4 - 0.721x^3 + 31.83x^2 - 594.6x + 4044.$ | 0.941 |
| **Missouri** | $y = 0.364x^4 - 32.67x^3 + 1095.x^2 - 16269x + 90434$ | 0.724 |
| **Arkansas** | $y = -1.558x^4 + 149.8x^3 - 5397.x^2 + 86389x - 51812$ | 0.254 |
| **Mississippi** | $y = 0.167x^4 - 15.47x^3 + 536.8x^2 - 8271.x + 47762$ | 0.642 |
| **South Dakota** | $y = 0.373x^4 - 28.01x^3 + 783.8x^2 - 9687.x + 44629$ | 0.775 |
| **Nebraska** | $y = -0.545x^4 + 39.36x^3 - 1061.x^2 + 12677x - 56531$ | 0.832 |
| **Delaware** | $y = 0.085x^4 - 7.010x^3 + 213.6x^2 - 2878.x + 14485$ | 0.583 |
| **United States <sup>a</sup>** | $y = -0.121x^4 + 8.709x^3 - 232.3x^2 + 2733.x - 11927$ | 0.869 |
| **PD vs Lower Endoscopy** |  |  |
|  | **Trendline Equation<sup>b</sup>** | **R-squared<sup>b</sup>** |
| **Maine** | $y = -0.001x^4 + 0.349x^3 - 36.45x^2 + 1680.x - 28873$ | 0.774 |
| **Vermont** | $y = -0.000x^4 + 0.181x^3 - 19.53x^2 + 925.2x - 16269$ | 0.822 |
| **Kansas** | $y = -0.000x^4 + 0.010x^3 - 1.302x^2 + 63.19x - 1070.$ | 0.970 |
| **Alaska** | $y = -0.002x^4 + 0.461x^3 - 40.66x^2 + 1584.x - 23025$ | 0.782 |
| **Missouri** | $y = -0.000x^4 + 0.197x^3 - 18.67x^2 + 779.5x - 12097$ | 0.961 |
| **Arkansas** | $y = -0.000x^4 + 0.068x^3 - 6.116x^2 + 238.4x - 3437.$ | 0.963 |
| **Mississippi** | $y = -0.000x^4 + 0.128x^3 - 10.98x^2 + 412.9x - 5756.$ | 0.965 |
| **South Dakota** | $y = 0.000x^4 - 0.077x^3 + 6.698x^2 - 257.9x + 3724.$ | 0.971 |
| **Nebraska** | $y = -0.000x^4 + 0.028x^3 - 2.611x^2 + 103.1x - 1490.$ | 0.957 |
| **Delaware** | $y = -0.008x^4 + 2.274x^3 - 241.7x^2 + 11382x - 20030$ | 0.759 |
| **United States <sup>a</sup>** | $y = 0.002x^4 - 0.563x^3 + 51.05x^2 - 2053.x + 30914$ | 0.994 |

Notes:
<sup>a</sup> All 50 states and the District of Columbia.
<sup>b</sup> Polynomial trendline equations and R-squared values were generated using Microsoft Excel.

As shown in Figures 1a-b and the tables, there is a inverse association between PD incidence and smoking for the U.S., but there is no association between PD and smoking for the ten states. As can be seen in Table 1, for PD incidence verses smoking, the R^2^ for the United States was 0.714 and ranged from a low of 0.004 for South Dakota to 0.613 for Mississippi. The average R^2^ for the ten states was 0.357.

As shown in Figures 2a-b and Table 1, there was no correlation between PD incidence and lower endoscopy utilization for the U.S. and for most of the ten states when the fitting curve examined in the regression analysis was linear. The R^2^ for the United States was 0.579 and ranged from a low of 0.349 for Delaware to 0.715 for Arkansas. The average R^2^ for the ten states was 0.588.

As can be seen from Tables 2-4, the best model fit to the data for PD verses lower endoscopy is a 3rd order polynomial. As shown in Table 3, the R^2^ for the United States was 0.971 and ranged from a low of 0.709 for Alaska to 0.970 for Kansas. The average R^2^ for the ten states was 0.878.

## DISCUSSION

The results presented here indicate that there is a strong positive correlation between PD incidence and lower endoscopy utilization. It seems likely that the inverse association between PD and smoking is confounded by lower endoscopy utilization.

The inverse association between PD and smoking is not seen in ages ≥75 years [16]; and the U.S. Preventive Services Task Force (USPSTF) recommends CRC screening for ages 50-75 years [17], further suggesting confounding by lower endoscopy utilization.

Lower endoscopy utilization reduces CRC risk [11-13]. Persons with PD have a decreased risk of CRC when compared to persons without PD [18-20], although this decreased risk is not seen in persons diagnosed with PD at an age of ≥75 years [18]. Since the USPSTF recommends CRC screening for ages 50-75 years [17], it seems possible that the inverse association between PD and CRC risk is also confounded by lower endoscopy utilization.

It may be that both the inverse association between PD and smoking and the inverse association between PD and CRC are confounded by the positive correlation between PD and lower endoscopy utilization. A biological explanation for a positive association between PD and lower endoscopy utilization is provided by the etiology of PD and suggests that iatrogenic transmission of PD pathology may be occurring.

A limitation of the present study is that the characteristics of the population base may not have remained unchanged throughout the study period. Other limitations include the smoking and lower endoscopy percentages provided by the CDC, which are estimates based on the Behavioral Risk Factor Surveillance System (BRFSS), and the PD incidence rates provided by the IHME, which are also estimates. Another limitation is that the column variable for current smoking was (_RFSMOK2) for 2004, but it was (_RFSMOK3) for all other years. Another limitation is that covariates of lower endoscopy utilization cannot be excluded.

## CONCLUSIONS

The present study concludes that there is a strong positive correlation between PD incidence and lower endoscopy utilization at a population level, reaffirming the findings of the previous study. The results suggest that the inverse association between PD and smoking is confounded by a positive association between PD and lower endoscopy utilization. Further investigation of the relationship between PD and lower endoscopy utilization is warranted and may provide a means for reducing PD incidence.

## Supporting information

strobe checklist

## Data Availability

PD incidence rates (age standardized, new cases per 100,000) are available from the Institute for Health Metrics and Evaluation (IHME)'s Global Burden of Diseases, Injuries, and Risk Factors (GBD) Compare visualization system (available at https://vizhub.healthdata.org/gbd-compare). Rates of current smoking (% adults who are current smokers) and rates of lower endoscopy utilization (% who ever had a sigmoidoscopy/colonoscopy) are available from the Centers for Disease Control and Prevention (CDC)'s Behavioral Risk Factor Surveillance System (BRFSS) Web Enabled Analysis Tool (WEAT) (available at https://www.cdc.gov/brfss/data_tools.htm).

## ACKNOWLEDGMENTS

The insight of Emily E. Davis was instrumental in the conception of this study.

